# Transmission of SARS-CoV-2 into and within immigrant households. Nation-wide registry-study from Norway

**DOI:** 10.1101/2021.07.09.21260253

**Authors:** Fredrik Methi, Rannveig Hart, Anna Godøy, Silje Jørgensen, Oliver Kacelnik, Kjetil Telle

## Abstract

**Background:** Minority ethnic groups and immigrants have been hit disproportionally hard by COVID-19 in many developed countries, including Norway. Most transmissions of SARS-CoV-2 occur in households.

**Methods:** Using individual-level registry data of all Norwegian residents we compared infections across all multi-person households. A household with at least one member born abroad was defined as an immigrant household. For the subset of households where at least one person tested positive for SARS-CoV-2 from August 1^st^ 2020 to May 1^st^ 2021, we calculated secondary attack rates (SARs) as the percent of other household members testing positive within 14 days after the first household member tested positive. Logistic regression model was used to adjust for sex, age, household composition and geography.

**Results:** Among all multi-person households in Norway (n=1 421 642), immigrant households (n=341 604) comprised more members on average (3.2) than households with only Norwegian-born members (2.8). The share of immigrant households where at least one member had been tested, was 56% (vs 49% in the households with only Norwegian-born members), and the share where at least one member was infected was 3.7% (vs 1.4% in households with only Norwegian-born members). Secondary attack rates were higher in immigrant (32%) than Norwegian-born households (20%). Results differed considerably by country of birth, with secondary attack rates particularly high in households from Syria, Iraq, Turkey, and Pakistan, also after adjustment for sex, age, household composition and geography.

**Conclusion:** SARS-CoV-2 is more frequently introduced into multi-person immigrant households than into households with only Norwegian-born members, and transmission within the household occurs more frequently in immigrant households. The results are likely related to living conditions, family composition or differences in social interaction, emphasizing the need to prevent introduction of SARS-CoV-2 into these vulnerable households.

## Introduction

Ethnic minority groups originating from West-Asia and Africa have been hit harder by COVID-19 than Ethnic Europeans in many European and North American countries [1-3]. Norway is no exception, as immigrants from Pakistan and Somalia, amongst others, have suffered more infections and hospitalizations than non-immigrants [4-5]. While some of the overrepresentation can be attributed to socioeconomic deprivation, substantial overrepresentation prevails after adjustment for demographic, socioeconomic, household and medical factors [1-6]. More knowledge is needed to design and implement measures that can break the chains of transmission.

Households are one of the most important arenas for transmission of SARS-CoV-2 [3, 7-8]. Little is known at the population level about characteristics of households more susceptible to infection and of households with more intra-household transmission, and this is particularly so among immigrants.

Using individual level data for all residents in Norway, our aim was to analyze transmissions of SARS-CoV-2 into and within all multi-person households in the country by the country or continent of birth of the household members.

## Methods

### Data

As part of the legally mandated responsibilities of The Norwegian Institute of Public Health (NIPH) during epidemics, a new emergency preparedness register, labelled BeredtC19, covering all residents of Norway, was established in cooperation with the Norwegian Directorate of Health in April 2020 [9]. The purpose of the preparedness register is to provide rapid overview and knowledge of how the pandemic and the measures that are implemented to contain the spread of the virus, affect the population’s health, use of health care services and health-related behaviors. The register contains daily updated information from the Norwegian Surveillance System for Communicable Diseases (MSIS), updated information from the Population Registry, and members of each household from Statistics Norway. All laboratories in Norway conducting polymerase chain reaction (PCR) tests for SARS-CoV-2 notify MSIS about the test result, the date of testing and the identity of the person tested. The time from sampling to results, which may influence secondary transmissions, has typically been less than one or two days [10]. Information was linked at the individual level using the unique personal identification number (encrypted version) provided to every Norwegian resident at birth or upon immigration.

We utilized the individual-level data in BeredtC19 for all residents of Norway, with vital demographic statistics (sex, year of birth, household members, etc.), and PCR tests for SARS-CoV-2 (dates, test result, etc.). Testing capacity was restricted before the summer of 2020, and we have thus limited the analysis to the period from August 1^st^ 2020 when testing was encouraged and free. Institutional board review was conducted, and the project was approved by the Ethics Committee of South-East Norway (March 9^th^, 2021, #198964).

### Population, definitions and time of follow up

Our population included all residents of Norway at the beginning of 2020 (5.4 million), implying that non-residents (like tourists, temporary workers and asylum applicants) were not available in the registry data. A household comprises every resident of one dwelling (excluding institutions), typically one family living in the same housing unit at the same address. We restricted the analyses to individuals who were living in households with at least two members (i.e. single-person households were excluded throughout the analysis).

A household with at least one member born abroad, was classified as an immigrant household. Immigrant households originating from countries with less than 300 infected households in our data, were attributed to the corresponding continent to allow for meaningful statistical analysis and ensure confidentiality. European countries were subdivided into three groups (‘Western Europe’, ‘Eastern Europe’ and ‘New EU-members’). The exact countries attributed to each group is shown in S-Table 6. To be able to make non-overlapping household definitions, we put immigrant households with members from more than one additional country of birth than Norway, in a separate category (‘more countries of origin’).

We focused on households with at least one member with a positive PCR-test for SARS-CoV-2 between August 1^st^ 2020 and May 1^st^ 2021, with follow-up time to May 15^th^ 2021. The first member of a household who tested positive in a PCR-test was defined as index case. Households where more members than the index case tested positive on the same day (“co-index”), were not included in calculations of secondary attack rates.

Secondary attack rates (SARs) were calculated by the share of household members (excluding the index case in both the numerator and denominator) with a positive test within 14 days after the sampling date of the index case (SAR14). SAR14 was calculated for the overall sample, as well as by the households’ country of origin.

### Analyses

We described characteristics of households with an index case and compared them to the overall population of multi-person households. Then we calculated the percent of households with an index case that had secondary infections, and calculated SAR14 for immigrant and non-immigrant households separately, both aggregated and by country of origin. To check that nearly all secondary cases were captured by 14 days, we also calculated SAR from 1 through 30 days after date of index case; to check that results were not driven by frequency of testing, we calculated the percent of household members who had been tested from 1 through 30 days after date of index case; and to check that results were not driven by variation in increasing vaccination during the spring of 2021, we calculated SAR14 by calendar month. Confidence intervals (95%) for SARs and percent of household members tested were calculated using the Wilson method. To explore robustness of SAR14 by country of origin to compositional differences (sex, age, household size, both young and old members in the same household, county of residence), we run a logistic regression model on the sample of all non-index household members, with secondary infection by 14 days as the outcome variable. All variables were included as categorical variables, i.e. sex (male/female), age (0-9, 10-19, 20-29, 30-39, 40-49, 50-59, 60-69, and 70 or above), household members (two, three, four, five, six, and seven or more), presence of both at least one household member below the age of 20 *and* at least one household member above the age of 60 (yes/no), and county of residence (the 11 administrative counties of Norway). The statistical software used was Stata MP v.16.

## Results

### Background statistics

Among multi-person households in Norway (n=1 421 642), immigrant households (n=341 604) comprised more members on average (3.2) than households with only Norwegian-born members (2.8); see S-Table 1. The percent of households that included members both below 20 and above 60 years of age was also higher (3.6% vs. 1.7%). Members of immigrant households were on average seven years younger (33) than households with only Norwegian-born members (40), and particularly so for households with members born in Eritrea (25), Syria (25), Somalia (25) and Afghanistan (26); see S-Table 1.

The share of households with at least one member hospitalized or dead from COVID-19 was higher in immigrant (3.7 households with hospitalized member per 1000, 1.7 households with dead member per 10 000) than Norwegian-born households (0.9 per 1000, 1.1 per 10 000) (S-Figure 1, S-Table 2). Pakistani households were among the most severely hit, with 20.5 per 1000 having at least one member hospitalized and 20.0 per 10 000 having at least one infected member. Also, households with members born in Somalia, Iraq, Turkey and Afghanistan were severely hit by COVID-19.

In 56% of immigrant households, at least one member had been tested, while this was the case in 49% of households with only Norwegian-born members. Testing was frequent in households from Pakistan, Iraq and Somalia. The share of *tested* immigrant households that were positive was 6.6%, while it was 2.8% in households with only Norwegian-born members (S-Table 1), suggesting higher infection rates in the immigrant populations (and not just that more tests revealed more infections).

### Transmissions into households

The share of *all* immigrant households (tested or not) with at least one member infected was 3.7%, compared with 1.4% in households with only Norwegian-born members; see S-Figure 1. There was substantial variation across country of origin, with at least one member having been positive in 12.9% of Pakistani households, in 10.3% of Somali households and in 9.5% of Iraqi households.

The high infection rates are not likely to be a result of more testing in immigrant households (cf. previous paragraph and S-Figure 1), as the share of the tested households that tested positive is also higher in immigrant households.

### Transmissions within households

Among the households with an index case, 42% of immigrant households and 29% of households with only Norwegian-born members had at least one secondary member infected by 14 days (S-Figure 2 and S-Table 3). After introduction of the virus into the household, more than 50% of households from Pakistan, Iraq and Eritrea had at least one more member infected within 14 days.

Secondary attack rates (SAR14) were also higher in immigrant (32%) than Norwegian-born households (20%); see Figure 1. Results differed considerably by country of birth, with the highest SAR14 in households from Syria (40%), Iraq (39%), Turkey (39%) and Pakistan (39%).

**Figure 1:**
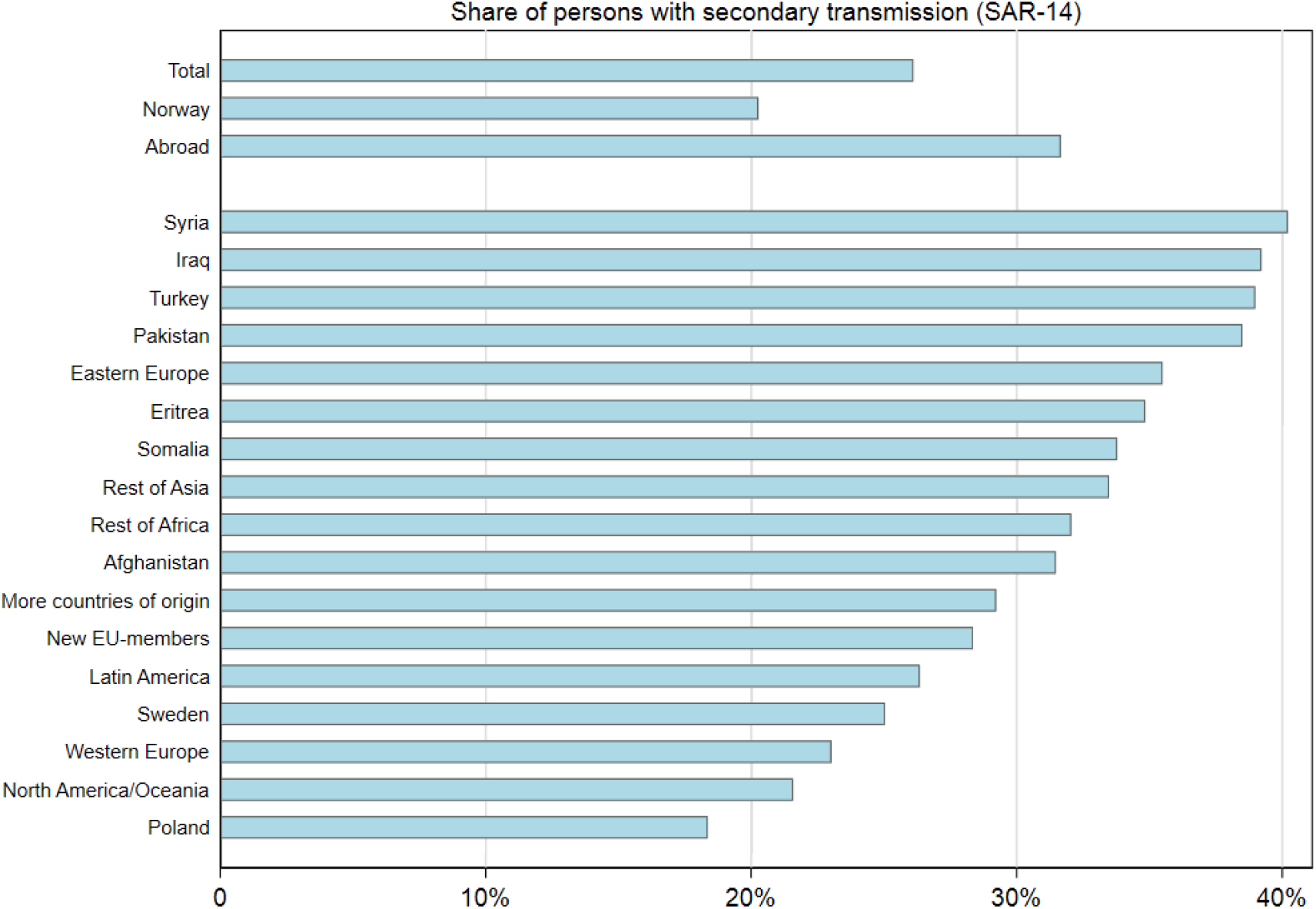
Secondary attack rates within 14 days after index sampling date (SAR14), by country of origin of household members. Note: All multi-person households in Norway, with the first household member (index case) testing positive for SARS-CoV-2 in a PCR test between August 1st 2020 and May 1st 2021. Exact rates and 95%CI are shown in Supplementary Table 4.

Results from the logistic regression model (Figure 2; S-Table 7) show that the elevated secondary transmissions by 14 days in immigrant households changed little when adjusting for age, sex and household size. When also adjusting for county of residence (Figure 2), the elevated secondary transmission fell somewhat for Iraq, Turkey and Pakistan, suggesting that households from these countries may have elevated SAR14 partly because they live in high-prevalence regions.

**Figure 2:**
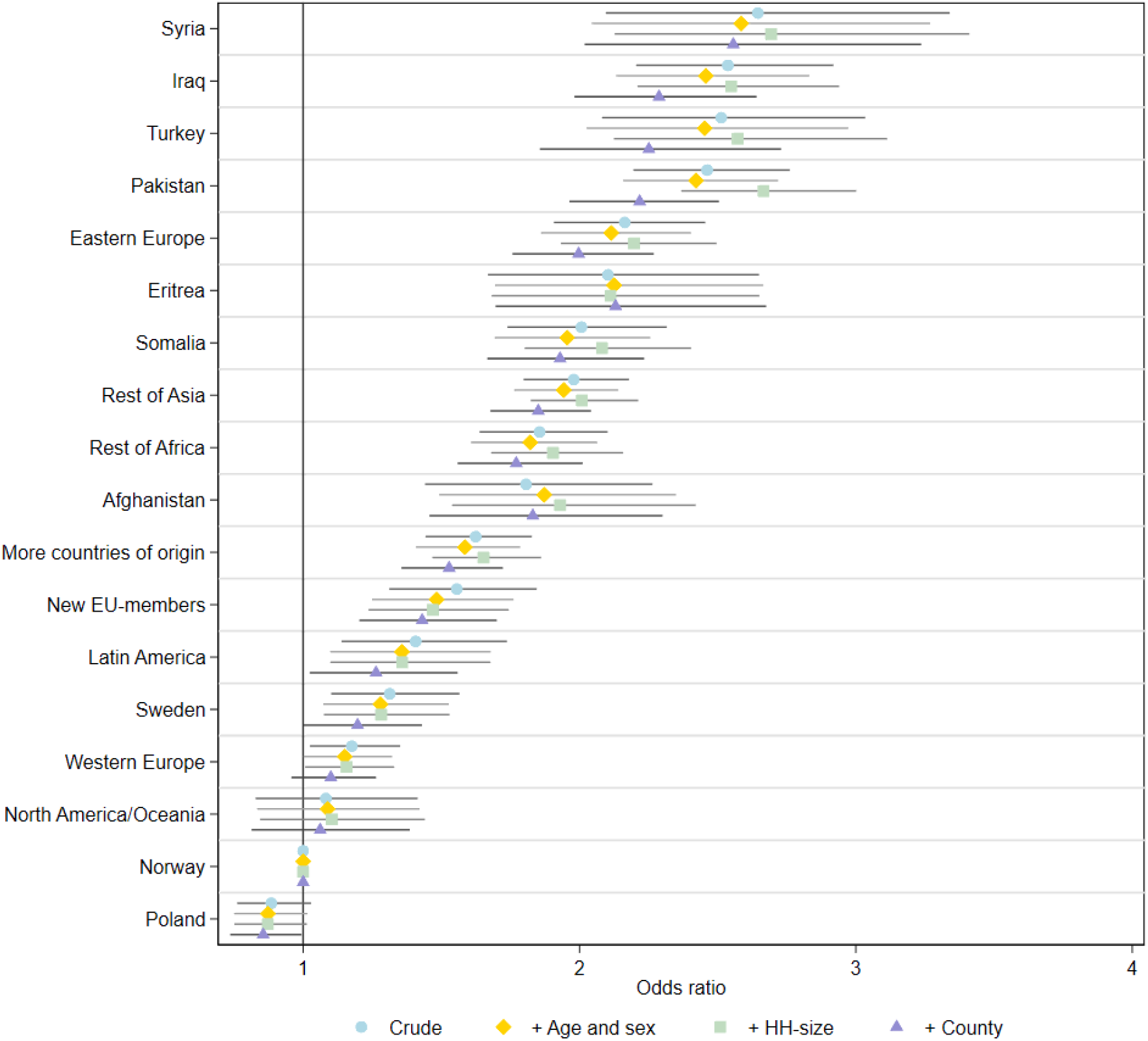
Logistic regression with secondary transmissions within 14 days after index sampling date as dependent variable, by household country of origin. Note: The first model (Crude) contains no control variables. The second model (+Age and sex) adjusts for age in ten-year intervals (0-9, 10-19, 20-29, 30-39, 40-49, 50-59, 60-69, 70+), and the person’s sex. The third model (+ HH-size) additionally adjusts for the number of persons in each household (two, three, four, five, six, seven or more) and whether the household contains persons both below 20 years and over 60 years of age. The fourth model (+County) additionally adjusts for the administrative county where the household is located. Coefficients are given in odds ratios with their respective 95% confidence intervals. Exact odds ratios and 95%CI are shown in Supplementary Table 7.

The share of household members tested within 14 days after the index case is high for all groups, although the testing rates in immigrant households were slightly below that of Norwegian-born households up to about nine days after index positivity, and thereafter higher (S-Figure 4). SAR14 tended to increase over calendar time, but it remained about ten percentage points higher in immigrant than Norwegian-born households in all calendar months (S-Figure 3; S-Table 5).

## Discussion

### Principal findings

Studying all residents in Norway living in multi-person households, we find that registered transmission of SARS-CoV-2 from August 1^st^ 2020 to May 1^st^ 2021 is substantially higher both into and within immigrant households than households with only Norwegian-born members.

An immigrant household was more than twice as likely to have a member infected with SARS-CoV-2 than households with only Norwegian-born members (3.7% vs 1.7%). Once a household member was infected, the secondary attack rate (SAR) by 14 days was more than 50 percent higher in immigrant households than in households with only Norwegian-born members (32% vs 20%). Results differed considerably by country of birth, with households from Syria, Iraq, Turkey, and Pakistan hit particularly hard. Main results were not altered by adjusting for age, sex, household composition and county of residence.

### Related studies

Though there are many studies showing higher infection rates among ethnic minority and immigrant groups originating from West-Asia and Africa also after adjusting for demographic, socioeconomic, household and medical factors [1-6], we are not aware of any previous study of transmissions into and within immigrant households. Previous studies of household SAR do not focus on differences in transmission across country of origin, and results on intra-household transmissions of SARS-CoV-2 are inconclusive, as the studies are few and small, with different designs, and report widely varying SARs [8, 11-18]. We observed an overall SAR in line with previous studies, e.g. in two systematic reviews Madewell *et al*. reported household SAR of 17% [11] and Lei *et al*. of 27% [13], neither providing information about ethnic groups or immigrants.

### Interpretations

Norway has based much of its pandemic response on a strategy of coordinated control measures. In the study period, this has included testing everyone after travel abroad and everyone with known exposure or minor symptoms, isolation of positive cases, careful contact tracing and quarantine and testing through the incubation period. The indications for testing, definitions of close contacts and length of quarantine have been regulated by law and adjusted over the course of the pandemic. Separating infected and non-infected household members by offering temporary alternative housing has only been done to a limited degree. In some periods, quarantine at a hotel has been mandatory for some categories of international travelers.

PCR-testing has been widely available in Norway, and from August 2020 anyone wanting a test could have one by contacting their local municipal test-station, where testing was free of charge. In our data the frequency of testing for immigrants has exceeded slightly that of Norwegian-born, but higher testing among immigrants is unlikely to drive our results as their positivity rate is also higher.

We found substantial differences in the share of infected households and in SARs by household country of origin. The twice as high transmission rate *into* immigrant households than households with only Norwegian-born members, could be related to the immigrant households being 15% larger (3.2 vs 2.8 members) with accompanying higher aggregated risk of one of the members being infected. Moreover, immigrants tend to live in urban areas that have had higher infection rates [5], which may also explain some of the higher introduction rates into these households. The government has prioritized vaccine distribution in Norway to areas with high infection rates and a high percentage of multi-person immigrant households. As this went into effect from March 2021, and were reinforced from June 2021, we may start to see declining differences in transmission into (and within) households by immigrant and non-immigrant descent.

Differences in social contact patterns, traveling and occupational risks have also been suggested as possible partial explanations [3-4, 19]. The recently published consensus statement from SAGE in the UK concludes that households are important drivers of COVID-19 transmission [3]. The paper is based on five published studies. All five studies found that household composition is crucial in understanding spread with a larger risk associated with larger multigenerational households. Whilst they support the idea that household composition may explain additional risk for some ethnic groups, they do not explain the total additional risk seen in some parts of the population.

We also found substantial differences in secondary transmissions *within* households by country of origin. This may be related to the composition of the households, and we observe, for example, that the share of immigrant households with members both below 20 and above 60 years of age is more than twice as high in immigrant households than in households with only Norwegian-born members (3.6% vs 1.7%). Nafilyan et al. [6] find that part of the ethnic inequalities in mortality can be explained by living in a multi-generational household, and Telle et al. [20] have shown that SAR is high from young children to their care givers. Also, older people may develop more symptoms and have a higher viral load, which may enhance intra-household spread. We observe households originating from the countries with highest intra-household SAR, are also those with most members and with both young and old members in the same household. However, in logistic regression models we find that the elevated secondary transmission in immigrant households persists after adjustment for household size and presence of both young and old members (Figure 2; S-Table 7).

Not only household size, but for example also housing space may contribute to the spread of the virus [4]. Norwegian advice has been that when isolating at home, cases should have as little contact with other family members as possible including where possible, their own bathroom and meals brought to them. This is harder in large families sharing smaller living spaces, and if young or old family members are dependent on close care from the others. In Norway, alternative housing to separate infected from non-infected household members has only been offered and accepted to a limited extent. Measures that make alternative housing more appealing, for instance moving the whole household to a larger dwelling rather than splitting out the index, may be considered.

Early testing of the index is important to break the chain of intra-household transmission, but so is also testing of the rest of the household. Small delays in such testing for possible secondary cases may affect intra-household secondary transmissions considerably [10]. Our data suggest that testing of immigrant households is somewhat delayed in the first week after index is positive (S-Figure 4), and the possible consequences of this deserves further research. We do not know why the testing is delayed, but possible reasons could be differences in health literacy and language difficulties.

The effect of strict travel control measures in preventing virus introduction into immigrant household is also worthy of more research. These should be contrasted with local measures aimed at, for example, increasing trust between immigrant communities and health authorities or restricting the size of social gatherings. Moreover, the municipal contact tracing teams should have access to professional interpreters in the communication with immigrant households.

One possible explanation for the higher intra-household secondary transmission in immigrant households compared to households with only Norwegian-born members, has been that the share of more easily transmittable virus variants is higher in these households. Between August 1^st^ 2020 and May 1^st^ 2021, the difference in SAR14 between immigrant and Norwegian-born households varied between 7 and 14 percentage points (S-Figure 3; S-Table 5). Before the first case of the Alpha variant was confirmed in Norway in December 2020 [21], the monthly difference varied between 9 and 12 percentage points. Hence, circulation of different variants is not a likely explanation for the observed differences.

### Potential limitations

We did not have data to confirm that the secondary cases were in fact transmissions from the index case. It is possible that both the index case and the secondary cases had a common external source, or that they were infected by different external sources. Better knowledge of actual directions of transmission within families would improve our ability to evaluate this, for example by judgements by health care personnel following each family or by genomic characterization of the viruses. However, several transmissions into the same household have been unlikely in Norway as the incidence rate of SARS-CoV-2 has been low throughout the pandemic. Still, immigrants more often live in urban areas with higher infection rates, possibly making several introduction events into immigrant households marginally more likely than for households with only Norwegian-born members.

A clear advantage of our registry-based study to most other studies, is that we do not have attrition: We observe every household, and we can observe all household members in the follow-up period, regardless of motivation to participate in a study or not. Indeed, our data stem from a real-world situation, where detection of secondary cases relates to a combination of the actual transmission of the virus and the behavioral responses to disease and the actual testing regime.

### Conclusions

By looking at register data of all Norwegian residents living in multi-person households, we see that households with immigrants are both more vulnerable to virus introduction into the household and to subsequent transmission within the household. More knowledge is needed to find the specific measures that can break the chains of transmissions into and within households, especially immigrant households that are hit particularly hard by COVID-19.

## Data Availability

Data are not publicly available.

## Acknowledgements

We would like to thank the Norwegian Directorate of Health, in particular Director for Health Registries Olav Isak Sjøflot and his department, for excellent cooperation in establishing the emergency preparedness register. We would also like to thank Gutorm Høgåsen and Anja Elsrud Schou Lindman for their invaluable efforts in the work on the register. The interpretation and reporting of the data are the sole responsibility of the authors, and no endorsement by the register is intended or should be inferred. We would also like to thank everyone at the Norwegian Institute of Public Health who has been part of the outbreak investigation and response team.

## Funding/support

The work was supported by the Norwegian Institute of Public Health. No external funding was received.

## Conflict of interest

All authors have completed the ICMJE uniform disclosure form and declare: no support from any organization for the submitted work; no financial relationships with any organizations that might have an interest in the submitted work in the previous three years; no other relationships or activities that could appear to have influenced the submitted work.

## Availability of data and material

Data are not publicly available.

## Code availability

STATA do-files are available upon request.

## Author contribution

Fredrik Methi had access to all of the data in the study and takes full responsibility for the integrity of the data and the accuracy of the data analysis. Rannveig Hart and Anna Godøy coded the initial dataset. Fredrik Methi and Kjetil Telle performed the statistical analyses and drafted the manuscript with Silje Jørgensen and Oliver Kacelnik. All authors contributed with acquisition of data, conceptual design, analyses and interpretation of results. All authors contributed to writing the article or critically revising it for important intellectual content. All authors gave final approval for the version to be submitted.

## Ethics approval

Institutional board review was conducted, and the project was approved by the Ethics Committee of South-East Norway (March 9^th^, 2021, #198964).

## Notes

### Competing Interest Statement

The authors have declared no competing interest.

### Author Declarations

Institutional board review was conducted, and the project was approved by the Ethics Committee of South-East Norway (March 9th, 2021, #198964).

