## Supplementary for "Transmission of SARS-CoV-2 into and within immigrant households. Nation-wide registry-study from Norway"

By Methi et al., 2021.


| | |
|---|---|
| Supplementary Table 1: Characteristics of all multi-person households | p. 2 |
| Supplementary Figure 1: Share of multi-person households | p. 3 |
| Supplementary Table 2: Shares of multi-person households | p. 4 |
| Supplementary Figure 2: Share of households with secondary transmission | p. 5 |
| Supplementary Table 3: Share of households with secondary transmission | p. 6 |
| Supplementary Table 4: Secondary attack rate (SAR14) | p. 7 |
| Supplementary Figure 3: SAR14 by month and country of origin | p. 8 |
| Supplementary Table 5: SAR14 by month and country of origin | p. 8 |
| Supplementary Figure 4: Cumulative proportion of SAR and tested | p. 9 |
| Supplementary Table 6: Classification of European countries | p. 9 |
| Supplementary Table 7: Logistic regression | p. 10-13 |

**Supplementary Table 1:** Characteristics of all multi-person households in Norway by country of birth of the household members. Number of households with at least one member who had been registered infected, tested, hospitalized or died because of COVID-19 between August 1st 2020 and May 1st 2021.

| Country | No of households | Mean HH size | Mean Age | HH with age < 20 and > 60 | HH with index | HH with tested | HH with hospitalized | HH with deaths | HH with secondary infection within 30 days | HH positive tests |
|---|---|---|---|---|---|---|---|---|---|---|
| Total | 1,421,642 | 2.9 | 38.3 | 30,233 (2.1%) | 27,669 (1.9%) | 721,293 (50.7%) | 2,246 (0.2%) | 176 (0.01%) | 9,627 (0.7%) | 3.8 % |
| Norway | 1,080,038 | 2.8 | 40.0 | 17,986 (1.7%) | 14,929 (1.4%) | 528,495 (48.9%) | 997 (0.1%) | 117 (0.01%) | 4,303 (0.4%) | 2.8 % |
| Abroad | 341,604 | 3.2 | 33.4 | 12,247 (3.6%) | 12,770 (3.7%) | 192,798 (56.4%) | 1,249 (0.4%) | 59 (0.02%) | 5,324 (1.6%) | 6.6 % |
| Rest of Asia | 66,539 | 3.2 | 34.6 | 4,231 (6.4%) | 1,908 (2.9%) | 35,186 (52.9%) | 249 (0.4%) | 15 (0.02%) | 835 (1.3%) | 5.4 % |
| Western Europe | 56,176 | 3.0 | 38.1 | 1,245 (2.2%) | 973 (1.7%) | 31,263 (55.7%) | 62 (0.1%) | 6 (0.01%) | 321 (0.7%) | 3.1 % |
| Poland | 29,421 | 2.9 | 33.8 | 256 (0.9%) | 1,064 (3.6%) | 15,639 (53.2%) | 50 (0.2%) | < 5 | 278 (0.9%) | 6.8 % |
| New EU-members | 27,227 | 2.9 | 31.6 | 392 (1.4%) | 554 (2.0%) | 12,403 (45.6%) | 24 (0.1%) | 0 | 214 (0.8%) | 4.5 % |
| More than one abroad | 27,180 | 3.4 | 30.7 | 1,178 (4.3%) | 1,244 (4.6%) | 16,232 (59.7%) | 120 (0.4%) | < 5 | 501 (1.8%) | 7.7 % |
| Sweden | 25,157 | 3.0 | 35.6 | 519 (2.1%) | 597 (2.4%) | 15,015 (59.7%) | 30 (0.1%) | 0 | 206 (0.8%) | 4.0 % |
| Eastern Europe | 21,841 | 3.2 | 34 | 892 (4.1%) | 984 (4.5%) | 12,281 (56.2%) | 120 (0.5%) | < 5 | 466 (2.1%) | 8.0 % |
| Rest of Africa | 17,635 | 3.4 | 29.8 | 879 (5.0%) | 1,031 (5.8%) | 10,962 (62.2%) | 108 (0.6%) | < 5 | 453 (2.6%) | 9.4 % |
| Latin America | 14,715 | 3.1 | 33.7 | 609 (4.1%) | 383 (2.6%) | 8,857 (60.2%) | 25 (0.2%) | < 5 | 141 (1.0%) | 4.3 % |
| North America/Oceania | 13,034 | 3.1 | 37.8 | 414 (3.2%) | 273 (2.1%) | 7,639 (58.6%) | 13 (0.1%) | < 5 | 81 (0.6%) | 3.6 % |
| Pakistan | 8,008 | 4.1 | 31.5 | 729 (9.1%) | 1,033 (12.9%) | 6,109 (76.3%) | 164 (2.0%) | 16 (0.20%) | 568 (7.1%) | 16.9 % |
| Somalia | 7,528 | 3.9 | 24.7 | 224 (3.0%) | 776 (10.3%) | 5,252 (69.8%) | 62 (0.8%) | < 5 | 330 (4.4%) | 14.8 % |
| Iraq | 6,854 | 3.7 | 28.8 | 233 (3.4%) | 648 (9.5%) | 4,692 (68.5%) | 88 (1.3%) | < 5 | 349 (5.1%) | 13.8 % |
| Syria | 5,876 | 3.8 | 25.4 | 115 (2.0%) | 310 (5.3%) | 3,191 (54.3%) | 32 (0.5%) | < 5 | 142 (2.4%) | 9.7 % |
| Eritrea | 5,428 | 3.2 | 25.3 | 69 (1.3%) | 309 (5.7%) | 2,621 (48.3%) | 18 (0.3%) | 0 | 122 (2.2%) | 11.8 % |
| Turkey | 4,937 | 3.5 | 32.0 | 151 (3.1%) | 382 (7.7%) | 2,956 (59.9%) | 46 (0.9%) | 6 (0.12%) | 199 (4.0%) | 12.9 % |
| Afghanistan | 4,048 | 3.5 | 26.4 | 111 (2.7%) | 301 (7.4%) | 2,497 (61.7%) | 38 (0.9%) | < 5 | 118 (2.9%) | 12.1 % |

**Supplementary Figure 1**: Share of multi-person households with at least one member who had been registered infected, tested, hospitalized or dead because of COVID-19 between August 1st 2020 and May 1st 2021.

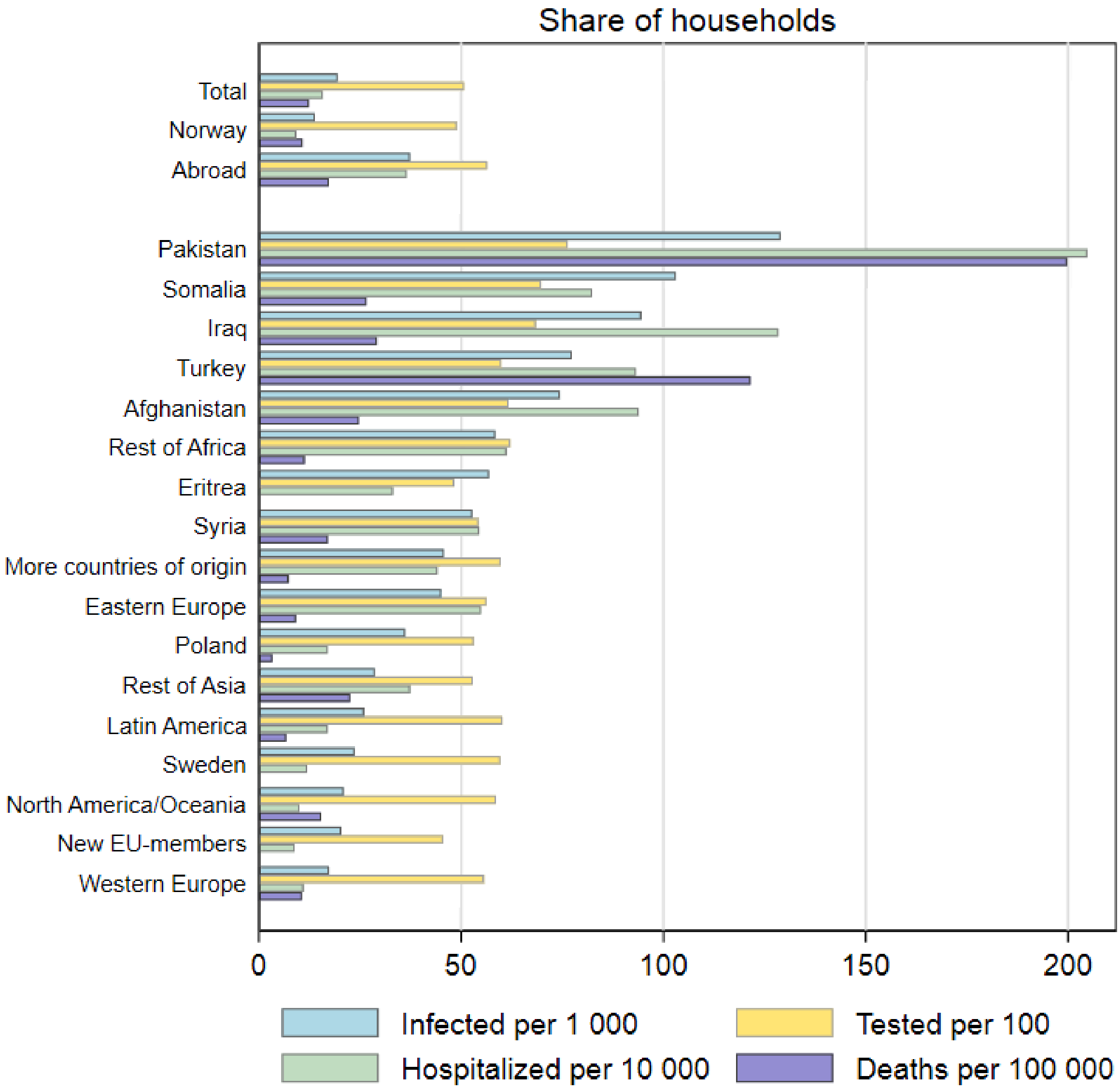

**Supplementary Table 2:** Shares of households with infections, tests, hospitalizations and deaths by country of origin.

| Country | Infections per 1000 | Tested per 100 | Hospitalizations per 10 000 | Deaths per 100 000 | Infections | Tests | Hospitals | Deaths | N |
|---|---|---|---|---|---|---|---|---|---|
| Total | 19.5 [19.5-19.7] | 50.7 [50.7-50.8] | 15.8 [15.2-16.5] | 12.4 [10.7-14.4] | 27699 | 721293 | 2246 | 176 | 1421642 |
| Norway | 13.8 [13.6-14.0] | 48.9 [48.8-49.0] | 9.2 [8.7-9.8] | 10.8 [9.0-13.0] | 14929 | 528495 | 997 | 117 | 1080038 |
| Abroad | 37.4 [36.8-38.0] | 56.4 [56.3-56.6] | 36.6 [34.6-38.6] | 17.3 [13.4-22.3] | 12770 | 192798 | 1249 | 59 | 341604 |
| Pakistan | 129.0 [121.8-136.5] | 73.6 [75.4-77.2] | 204.8 [176.0-238.2] | 199.8 [123.0-324.3] | 1033 | 6109 | 164 | 16 | 8008 |
| Somalia | 103.1 [96.4-110.2] | 69.8 [68.7-70.8] | 82.4 [64.3-105.4] | - | 776 | 5252 | 62 | < 5 | 7528 |
| Iraq | 94.5 [87.8-101.7] | 68.5 [67.3-69.5] | 128.4 [104.3-157.9] | - | 648 | 4692 | 88 | < 5 | 6854 |
| Turkey | 77.4 [70.2-85.2] | 59.9 [58.5-61.2] | 93.2 [69.9-124.0] | 121.5 [55.7-264.9] | 382 | 2956 | 46 | 6 | 4937 |
| Afghanistan | 74.4 [66.7-82.8] | 61.7 [60.2-63.2] | 93.9 [68.4-128.6] | 24.7 [4.4-139.8] | 301 | 2497 | 62 | 6 | 4,048 |
| Rest of Africa | 58.5 [55.1-62.0] | 62.2 [61.4-62.9] | 61.2 [50.8-73.9] | - | 1031 | 10962 | 108 | < 5 | 17635 |
| Eritrea | 56.9 [51.1-63.4] | 48.3 [47.0-49.6] | 33.2 [21.0-52.4] | 0 | 309 | 2621 | 18 | 0 | 5428 |
| Syria | 52.8 [47.3-58.8] | 54.3 [53.0-55.6] | 54.5 [38.6-76.8] | - | 310 | 3191 | 32 | < 5 | 5876 |
| More countries of origin | 45.8 [43.3-48.3] | 59.7 [59.1-60.3] | 44.2 [36.9-52.8 | - | 1244 | 16232 | 120 | < 5 | 27180 |
| Eastern Europe | 45.1 [42.4-47.9] | 56.2 [55.6-56.9] | 54.9 [46.0-65.7] | - | 984 | 12281 | 120 | < 5 | 21841 |
| Poland | 36.2 [34.1-38.4] | 53.2 [52.6-53.7] | 17.0 [12.9-22.4] | - | 1064 | 15639 | 50 | < 5 | 29421 |
| Rest of Asia | 28.7 [27.4-30.0] | 52.9 [52.5-53.3] | 37.4 [33.1-42.4] | 22.5 [13.7-37.2] | 1908 | 35186 | 249 | 15 | 66539 |
| Latin America | 26.0 [23.6-28.7] | 60.2 [59.4-61.0] | 17.0 [11.5-25.1] | - | 383 | 8857 | 25 | < 5 | 14715 |
| Sweden | 23.7 [21.9-25.7] | 59.7 [59.1-60.3] | 11.9 [8.4-17.0] | 0 | 597 | 15015 | 30 | 0 | 25157 |
| North America/Oceania | 20.9 [18.6-23.5] | 58.6 [57.7-59.5] | 10.0 [5.8-17.1] | - | 273 | 7639 | 13 | < 5 | 13034 |
| New EU-members | 20.3 [18.7-22.1] | 45.6 [45.0-46.1] | 8.8 [5.9-13.1] | - | 554 | 12403 | 24 | 0 | 27227 |
| Western Europe | 17.3 [16.3-18.4] | 55.7 [55.2-56.1] | 11 [8.6-14.1] | 10.7 [4.9-23.3] | 973 | 31263 | 62 | 6 | 56176 |

Note: 95% CIs around the estimated rates were calculated using the Wilson method.

**Supplementary Figure 2:** Share of households where at least one member tested positive by 14 days after the testing date of the index case. For households where index case tested positive between August 1st 2020 and May 1st 2021.

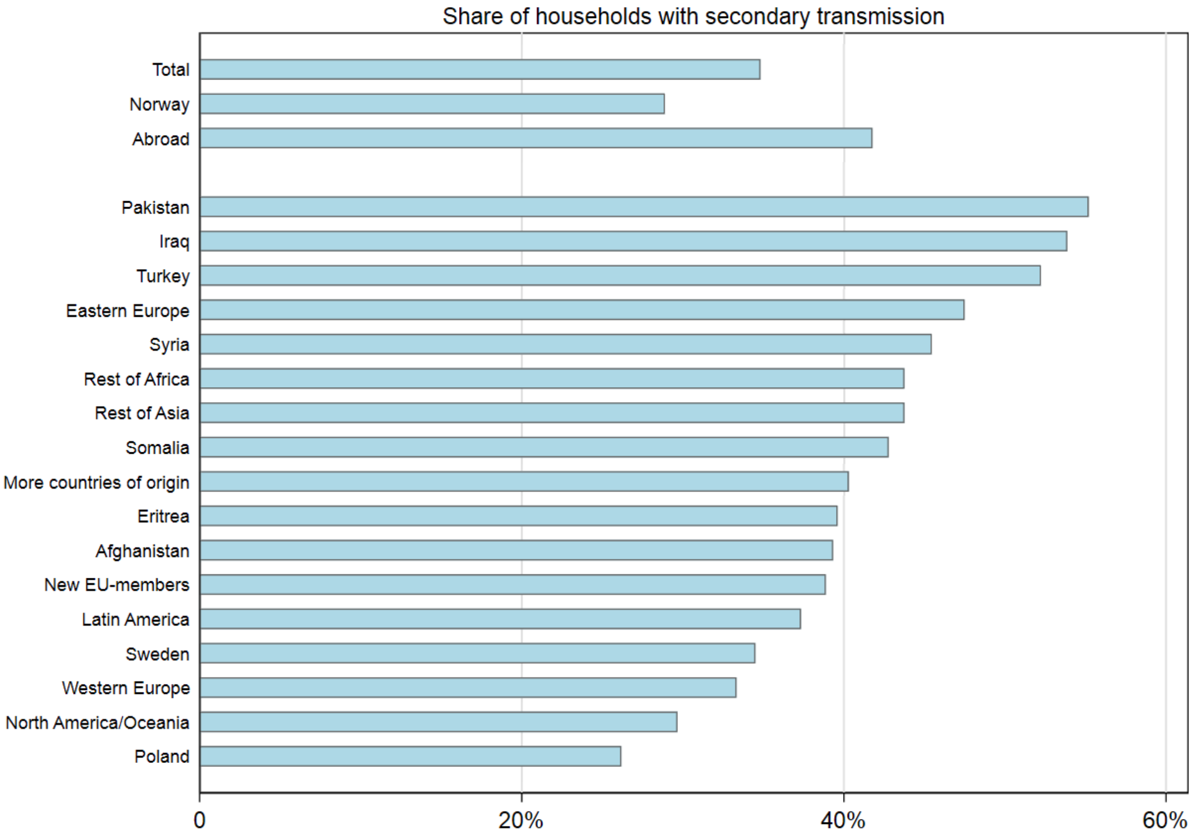

**Supplementary Table 3:** Share of households where at least one member tested positive by 14 days after the testing date of the index case. For households where index case tested positive between August 1st 2020 and May 1st 2021.

| Country | Share of households with secondary transmission | HH with secondary transmission | N |
|---|---|---|---|
| Total | 0.35 [0.34-0.35] | 9,628 | 27,650 |
| Norway | 0.29 [0.28-0.30] | 4,308 | 14,915 |
| Abroad | 0.42 [0.41-0.43] | 5,320 | 12,735 |
| Pakistan | 0.55 [0.52-0.58] | 568 | 1,029 |
| Iraq | 0.54 [0.50-0.58] | 348 | 646 |
| Turkey | 0.52 [0.47-0.57] | 199 | 381 |
| Eastern Europe | 0.47 [0.44-0.51] | 465 | 979 |
| Syria | 0.45 [0.40-0.51] | 140 | 308 |
| Rest of Africa | 0.44 [0.41-0.47] | 449 | 1,026 |
| Rest of Asia | 0.44 [0.42-0.46] | 831 | 1,899 |
| Somalia | 0.43 [0.39-0.46] | 332 | 776 |
| More countries of origin | 0.40 [0.38-0.43] | 501 | 1,243 |
| Eritrea | 0.40 [0.34-0.45] | 122 | 308 |
| Afghanistan | 0.39 [0.34-0.45] | 118 | 300 |
| New EU-members | 0.39 [0.35-0.43] | 215 | 553 |
| Latin America | 0.37 [0.33-0.42] | 143 | 383 |
| Sweden | 0.35 [0.31-0.38] | 206 | 597 |
| Western Europe | 0.33 [0.30-0.36] | 324 | 972 |
| North America/Oceania | 0.30 [0.25-0.35] | 81 | 273 |
| Poland | 0.26 [0.24-0.29] | 278 | 1,062 |

Note: 95% CIs around the estimated rates were calculated using the Wilson method.

**Supplementary Table 4:** Secondary attack rate (SAR14) was calculated as the number of non-index household members who tested positive within 14 days after the date when the index household member tested positive, divided by all non-index household members.

| Country | SAR-14 | Persons with secondary transmission | N |
|---|---|---|---|
| Total | 0.26 [0.26-0.26] | 17,584 | 67,343 |
| Norway | 0.20 [0.20-0.21] | 6,657 | 32,835 |
| Abroad | 0.32 [0.31-0.32] | 10,927 | 34,508 |
| Syria | 0.40 [0.37-0.43] | 372 | 925 |
| Iraq | 0.39 [0.37-0.41] | 769 | 1,961 |
| Turkey | 0.39 [0.36-0.42] | 439 | 1,126 |
| Pakistan | 0.39 [0.37-0.40] | 1,403 | 3,644 |
| Eastern Europe | 0.35 [0.34-0.37] | 952 | 2,682 |
| Eritrea | 0.35 [0.31-0.39] | 231 | 663 |
| Somalia | 0.34 [0.32-0.36] | 864 | 2,557 |
| Rest of Asia | 0.33 [0.32-0.35] | 4,821 | 1,614 |
| Rest of Africa | 0.32 [0.30-0.34] | 940 | 2,932 |
| Afghanistan | 0.31 [0.28-0.35] | 260 | 826 |
| More countries of origin | 0.29 [0.28-0.31] | 1,096 | 3,750 |
| New EU-members | 0.28 [0.26-0.31] | 328 | 1,157 |
| Latin America | 0.26 [0.24-0.29] | 229 | 869 |
| Sweden | 0.25 [0.23-0.27] | 346 | 1,382 |
| Western Europe | 0.23 [0.21-0.25] | 521 | 2,263 |
| North America/Oceania | 0.22 [0.19-0.25] | 142 | 658 |
| Poland | 0.18 [0.17-0.20] | 421 | 2,292 |

Note: 95% CIs around the estimated rates were calculated using the Wilson method.

**Supplementary Figure 3:** SAR14 by month and country of origin between August 1st 2020 and May 1st 2021.

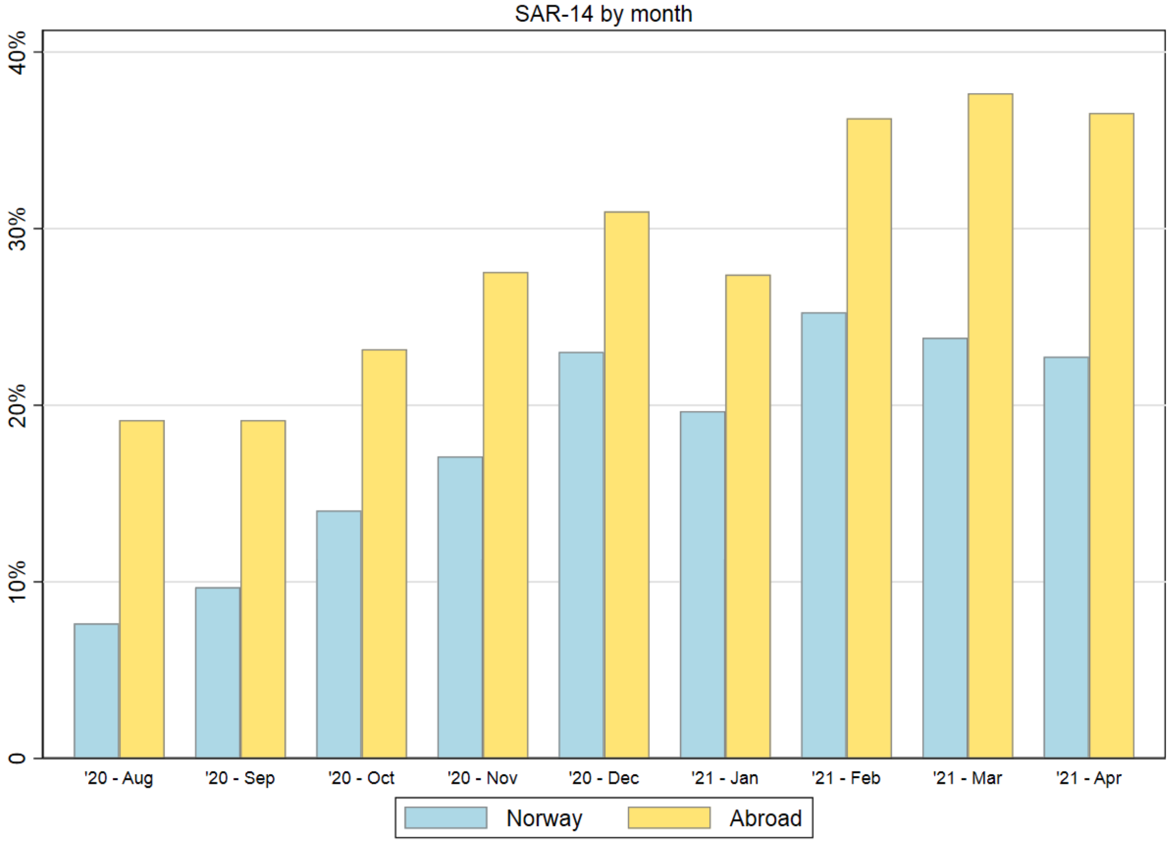


**Supplementary Table 5:** SAR14 by month and country of origin between August 1st 2020 and May 1st 2021.

| Month | Norway | Abroad |
|---|---|---|
| August 2020 | 7% (6%-10%) | 19% (16%-23%) |
| September 2020 | 10% (8%-11%) | 19% (17%-21%) |
| October 2020 | 14% (13%-15%) | 23% (22%-25%) |
| November 2020 | 17% (16%-18%) | 28% (26%-29%) |
| December 2020 | 23% (22%-24%) | 31% (30%-32%) |
| January 2021 | 20% (18%-21%) | 27% (26%-29%) |
| February 2021 | 25% (24%-27%) | 36% (35%-38%) |
| March 2021 | 24% (23%-25%) | 38% (37%-39%) |
| April 2021 | 23% (22%-24%) | 37% (35%-38%) |

Note: 95% CIs around the estimated rates were calculated using the Wilson method.

**Supplementary Figure 4:** Cumulative proportion of secondary transmission and members tested within households by household country of origin, for households where index case tested positive between August 1st 2020 and May 1st 2021

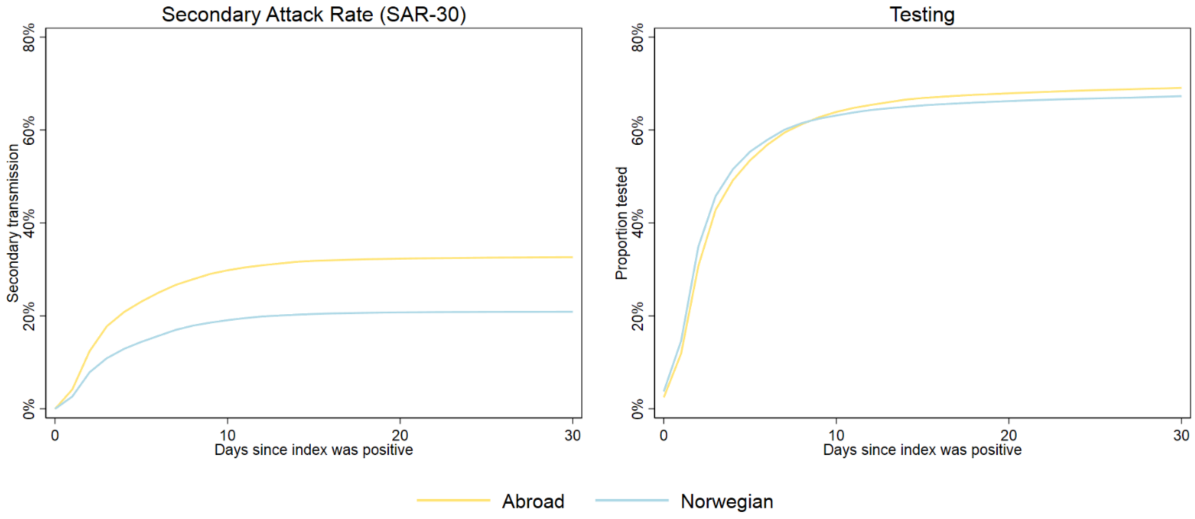


**Supplementary Table 6:** Classification of European countries:

| **Western Europe** | **New EU-members** | **Eastern Europe** |
|---|---|---|
| Andorra | Bulgaria | Albania |
| Austria | Croatia | Belarus |
| Belgium | Czech Republic | Bosnia-Herzegovina |
| Denmark | Cyprus | Kosovo |
| Faeroe Island | Estonia | Moldova |
| Finland | Hungary | Montenegro |
| France | Latvia | North-Macedonia |
| Germany | Latvia | Russia |
| Gibraltar | Lithuania | Serbia |
| Greece | Romania | Ukraine |
| Greenland | Slovakia | |
| Iceland | | |
| Ireland | | |
| Italy | | |
| Liechtenstein | | |
| Luxembourg | | |
| Monaco | | |
| Netherlands | | |
| Portugal | | |
| San Marino | | |
| Spain | | |
| Switzerland | | |
| United Kingdom | | |

Note: Sweden and Poland are analyzed individually because they constitute large groups.

**Supplementary Table 7:** Logistic regression for secondary attack rates within 14 days after index sampling date (SAR14), by country of origin of household members.

| | (1) Crude | (2) + Age and sex | (3) + HH-size | (4) + County |
|---|---|---|---|---|
| Country | | | | |
| Abroad | 1.822*** | 1.782*** | 1.806*** | 1.648*** |
| | [1.733-1.916] | [1.693-1.875] | [1.717-1.900] | [1.565-1.737] |
| Afghanistan | 1.806*** | 1.872*** | 1.930*** | 1.831*** |
| | [1.441-2.264] | [1.492-2.349] | [1.539-2.421] | [1.457-2.301] |
| Eastern Europe | 2.164*** | 2.115*** | 2.196*** | 1.997*** |
| | [1.907-2.455] | [1.861-2.403] | [1.933-2.496] | [1.757-2.269] |
| Eritrea | 2.103*** | 2.125*** | 2.111*** | 2.130*** |
| | [1.669-2.650] | [1.694-2.665] | [1.682-2.651] | [1.696-2.675] |
| Iraq | 2.537*** | 2.457*** | 2.549*** | 2.288*** |
| | [2.205-2.919] | [2.132-2.831] | [2.210-2.939] | [1.982-2.641] |
| Latin America | 1.407** | 1.357** | 1.358** | 1.264* |
| | [1.139-1.738] | [1.098-1.678] | [1.098-1.678] | [1.024-1.559] |
| More countries of origin | 1.624*** | 1.585*** | 1.653*** | 1.528*** |
| | [1.443-1.828] | [1.407-1.785] | [1.468-1.861] | [1.355-1.723] |
| New EU-members | 1.556*** | 1.483*** | 1.469*** | 1.431*** |
| | [1.312-1.846] | [1.249-1.761] | [1.237-1.744] | [1.204-1.700] |
| North America/Oceania | 1.082 | 1.088 | 1.103 | 1.062 |
| | [0.828-1.414] | [0.833-1.421] | [0.844-1.440] | [0.813-1.386] |
| Norway (Ref.) | 1 | 1 | 1 | 1 |
| | [1-1] | [1-1] | [1-1] | [1-1] |
| Pakistan | 2.462*** | 2.422*** | 2.666*** | 2.218*** |
| | [2.195-2.761] | [2.157-2.719] | [2.368-3.001] | [1.963-2.505] |
| Poland | 0.885 | 0.873 | 0.872 | 0.855* |
| | [0.761-1.029] | [0.750-1.015] | [0.750-1.013] | [0.736-0.994] |
| Rest of Africa | 1.856*** | 1.822*** | 1.904*** | 1.771*** |
| | [1.638-2.102] | [1.608-2.064] | [1.680-2.158] | [1.559-2.012] |
| Rest of Asia | 1.979*** | 1.943*** | 2.009*** | 1.851*** |
| | [1.798-2.179] | [1.764-2.141] | [1.824-2.213] | [1.678-2.042] |

| | | | | |
|---|---|---|---|---|
| Somalia | 2.007*** | 1.955*** | 2.081*** | 1.929*** |
| | [1.739-2.316] | [1.694-2.256] | [1.802-2.404] | [1.667-2.234] |
| Sweden | 1.313** | 1.280** | 1.282** | 1.197* |
| | [1.102-1.566] | [1.072-1.527] | [1.075-1.530] | [1.002-1.430] |
| Syria | 2.645*** | 2.585*** | 2.694*** | 2.556*** |
| | [2.095-3.339] | [2.044-3.269] | [2.127-3.410] | [2.019-3.237] |
| Turkey | 2.513*** | 2.453*** | 2.572*** | 2.251*** |
| | [2.081-3.034] | [2.025-2.972] | [2.125-3.113] | [1.857-2.729] |
| Western Europe | 1.176* | 1.150* | 1.157* | 1.1 |
| | [1.024-1.351] | [1.001-1.322] | [1.007-1.329] | [0.958-1.263] |
| Age | | | | |
| 0-9 years (Ref.) | | 1 | 1 | 1 |
| | | | [1-1] | [1-1] |
| 10-19 years | | 1.057 | 1.067* | 1.089** |
| | | [0.993-1.125] | [1.003-1.135] | [1.023-1.159] |
| 20-29 years | | 0.518*** | 0.497*** | 0.522*** |
| | | [0.480-0.557] | [0.461-0.536] | [0.484-0.563] |
| 30-39 years | | 1.028 | 0.986 | 1.002 |
| | | [0.966-1.094] | [0.926-1.050] | [0.941-1.068] |
| 40-49 years | | 0.979 | 0.965 | 0.985 |
| | | [0.919-1.044] | [0.906-1.029] | [0.923-1.050] |
| 50-59 years | | 0.738*** | 0.715*** | 0.744*** |
| | | [0.686-0.794] | [0.665-0.769] | [0.692-0.801] |
| 60-69 years | | 0.919 | 0.887* | 0.922 |
| | | [0.831-1.015] | [0.803-0.980] | [0.834-1.019] |
| 70+ years | | 1.067 | 1.007 | 1.055 |
| | | [0.932-1.220] | [0.880-1.153] | [0.920-1.211] |
| Sex | | | | |
| Female (Ref.) | | 1 | 1 | 1 |
| | | [1-1] | [1-1] | [1-1] |
| Male | | 0.877*** | 0.879*** | 0.880*** |

| | | | |
|---|---|---|---|
| | [0.848-0.907] | [0.850-0.909] | [0.851-0.910] |
| Household size | | | |
| 2 (Ref.) | | 1 | 1 |
| | | [1-1] | [1-1] |
| 3 | | 0.833*** | 0.844*** |
| | | [0.775-0.895] | [0.786-0.908] |
| 4 | | 0.840*** | 0.854*** |
| | | [0.784-0.900] | [0.797-0.916] |
| 5 | | 0.756*** | 0.782*** |
| | | [0.695-0.822] | [0.719-0.851] |
| 6 | | 0.865* | 0.9 |
| | | [0.771-0.971] | [0.801-1.012] |
| 7+ | | 0.699*** | 0.753*** |
| | | [0.604-0.808] | [0.651-0.872] |
| Non-generational | | 1 | 1 |
| | | [1-1] | [1-1] |
| Generational | | 0.779** | 0.765*** |
| | | [0.672-0.904] | [0.660-0.888] |
| County | | | |
| Oslo (Ref.) | | | 1 |
| | | | [1-1] |
| Rogaland | | | 0.412*** |
| | | | [0.327-0.519] |
| Møre og Romsdal | | | 0.0565*** |
| | | | [0.0182-0.176] |
| Nordland | | | 0.846 |
| | | | [0.684-1.046] |
| Viken | | | 0.898*** |
| | | | [0.844-0.956] |
| Innlandet | | | 0.261*** |
| | | | [0.182-0.375] |

| | | | | | |
|---|---|---|---|---|---|
| | Vestfold og Telemark | | | | 0.843*** |
| | | | | | [0.764-0.930] |
| | Agder | | | | 0.806** |
| | | | | | [0.708-0.917] |
| | Vestland | | | | 0.650*** |
| | | | | | [0.591-0.714] |
| | Trøndelag | | | | 0.156*** |
| | | | | | [0.104-0.235] |
| | Troms og Finnmark | | | | 0.432*** |
| | | | | | [0.345-0.541] |
| | Observations | 67 343 | 67 343 | 67 343 | 67 343 |

Note: The first model (Crude) contains no control variables. The second model (+Age and sex) adjusts for age in ten year intervals (0-9, 10-19, 20-29, 30-39, 40-49, 50-59, 60-69, 70+), and the person's sex. The third model (+HH-size) additionally adjusts for the number of persons in each household (two, three, four, five, size, seven or more) and whether the household contains persons both below 20 years and over 60 years old. The fourth model (+County) additionally adjusts for the administrative county where the household is located. Coefficients are given in odds ratios with their respective 95% confidence intervals in brackets.

* p< 0.05, ** p<0.01, *** p<0.001
